# Healthcare Equity in Epilepsy Surgery: Equivalent Outcomes Between NAEC-IV Public Safety-Net and Tertiary Academic Centers in a Major Metropolitan Area

**DOI:** 10.1101/2024.11.04.24316317

**Authors:** Jordan Lam, Vivek Mehta, Jonathan Russin, David Millett, Susan Shaw, Lauren Liu, Brian Lee, Laura Kalayjian, Michelle Armacost, Hui Gong, Christi Heck, Darrin Lee, Charles Liu

## Abstract

Access to and delivery of quality surgical epilepsy care remains a challenge in both safety-net and private hospital systems. Underserved populations are more likely to utilize safety-net hospital systems, but there are few data on epilepsy surgery outcomes in this setting. We aimed to analyze and compare surgical epilepsy care in a safety-net versus private hospital system. We prospectively collected evaluation and treatment data in patients undergoing resective surgery for epilepsy at a safety-net hospital system and a collaborating private hospital system between 2010-2017. Seizure characteristics, pre-surgical evaluation, perioperative complications, and seizure outcomes were prospectively recorded. Data from 102 patients in the safety-net and 145 patients in the private hospital system were analyzed. There were higher proportions of African American (p=.02) and Hispanic patients (p= .03) in the safety-net hospital system. There was no difference in mean time from epilepsy onset to surgery between groups (p=.54). The presurgical evaluation was equivalent (p>.18), except for more frequent use of magnetoencephalography in the private system (p=.02). Seizure freedom outcomes were excellent, and complication rates were low with no significant differences between groups (p=.95 and p>.22, respectively). However, patients from safety-net hospital systems were more likely to be lost to follow up (p=.04). Quality and equitable surgical epilepsy care can be delivered in a safety-net hospital system despite the higher-minority demographics and intrinsic factors of safety-net hospitals. Partnership of safety-net hospital systems with established comprehensive epilepsy centers and the expansion of these services is essential for healthcare equity in modern epilepsy care.

## Introduction

The role for epilepsy surgery in the appropriate patient with medically refractory epilepsy is supported by strong class 1 data (1). In these patients, surgery is cost effective (2), superior to best medical management (1), improves overall quality of life (3), and reduces mortality (4-6). Access to a comprehensive epilepsy center with surgical capabilities is the largest barrier in the delivery of standard of care treatment in this medically refractory population. Numerous studies have documented factors such as socioeconomic status, racial background, language, and other disparities leading to healthcare inequity in surgical epilepsy care (1). These underserved patients are more likely to obtain care from safety-net hospitals. which the Institute of Medicine defines as those which 1) organize and deliver a significant level of health care to patients with no insurance or with Medicaid; 2) who play a major role in the provision of comprehensive services to medically and socially vulnerable populations; and 3) are providers of last resort (7). Recent reports on safety-net hospital systems have found higher illness severity, higher surgical complication rates, higher cost of care and inferior performance raising concerns about the delivery of surgical epilepsy care in this setting (8-10). To date the outcomes of epilepsy surgery in safety-net versus private hospital systems has not been compared.

In this study, we prospectively collected data on the delivery of surgical epilepsy care in a safety-net hospital system and a private hospital system. We evaluated for differences in baseline patient characteristics, presurgical evaluation, surgical morbidity and mortality and post-surgical seizure freedom and control outcomes. We describe the unique challenges and opportunities for providing quality surgical epilepsy care and describe strategies for developing these programs.

## Methods

### Study Population

We prospectively collected data from 102 patients with medically refractory epilepsy who underwent epilepsy surgery between 1/1/2010 and 1/1/2017 at the Los Angeles County Department of Health Services Comprehensive Epilepsy Center and 145 patients who underwent epilepsy surgery between 2009 and 2016 at the University of Southern California (USC) Comprehensive Epilepsy Center of the Keck Medical Center of USC. We prospectively collected data regarding baseline epilepsy characteristics, pre-surgical evaluation, surgical complications, and post-surgery seizure free outcomes. Institutional review board approval was obtained from the Rancho Research Institute and the University of Southern California.

### Pre-Surgical Evaluation

All patients were evaluated with a detailed history and physical exam and all previous electroencephalogram (EEG) monitoring, medication trials and surgery were reviewed. Patients were defined as refractory after a documented trial of two appropriate and effective doses of antiepileptic medications without seizure freedom. All refractory patients underwent routine presurgical evaluation including phase 1 long-term scalp video EEG monitoring, neuropsychological testing and trial of appropriate anti-epileptic medication. All patients underwent high-resolution magnetic resonance imaging (MRI) with thin cut coronal reconstructions for evaluation of the hippocampi. MRI images were obtained on a 1.5 or 3T MRI scanner and included coronal and oblique sequences. Positron emission tomography (PET) imaging was performed if phase 1 or MRI data were discordant, if there were distinct semiolgies or if it was believed that additional PET data could better identify a focus and aid with phase 2 coverage planning. Magnetoencephalography (MEG) was available at the private hospital system but not the safety-net hospital system. All patients were discussed in the same consensus multidisciplinary epilepsy conference with regards to invasive workup and treatment management.

### Surgery

Phase 2 invasive EEG monitoring was performed with either limited mesial coverage (subtemporal strips or depth electrodes), complete temporal coverage (lateral temporal cortex grids and subtemporal strips) or tailored coverage with combination of strips, grids or depth electrodes as deemed necessary for a period of time to capture the medically refractory seizures. The decision to perform invasive phase 2 monitoring was made when there was discordance with semiology, imaging, and/or phase 1 monitoring findings. Resective surgery consisted of either standard anterior temporal lobectomy or selective amygdalohippocampectomy with or without resection of the temporal pole or focal tailored resection with or without electrocorticography. The decision to perform surgery was based on a consensus determined at a multi-disciplinary epilepsy conference consisting of epileptologists, neurosurgeons, neuropsychologists, and neuroradiologists.

### Data Collection

The baseline clinical epilepsy characteristics, presurgical evaluation, surgical complications, and post-surgery seizure free outcomes were documented in the patient’s electronic medical record and were also recorded in a data management system.

### Outcomes

Surgical complications were recorded as follows: unanticipated neurological deficit, wound infection, symptomatic intracranial hemorrhage, stroke, reoperation within 24 hours for any reason, misplacement of monitoring leads, and subtotal resection of epileptogenic focus.

The following pertinent perioperative complications related to surgery were recorded: deep vein thrombosis (DVT), pulmonary embolus (PE), pneumonia, urinary tract infection (UTI), pressure ulcer, complications of anesthesia or death. Postoperative seizure outcomes were classified utilizing the Engel Epilepsy Surgery Outcome Scale (11) as follows: class I: free of disabling seizures; class II: rare disabling seizures (“ almost seizure-free”); class III: worthwhile improvement; Class IV: no worthwhile improvement. Outcomes data were collected at the immediate two-week and one-month post-operative visits in the combined neurosurgery and neurology epilepsy clinic and then with every subsequent clinic visit with neurology or neurosurgery. All patients continued on their dose of pre-surgical antiepileptic medication after surgery and were subsequently tapered off by the epilepsy neurologist.

### Statistical analysis

Baseline clinical characteristics (patient’s age, gender, seizure duration, risk factors), pre-surgical evaluation, surgical morbidity and mortality, and seizure free outcomes were compared between the two groups with the Pearson chi-square or Fisher’s exact test (nominal/ordinal), Student’s t-test (parametric continuous) or Mann-Whitney U test (nonparametric continuous) where appropriate. A p-value of <.05 was considered statistically significant.

## Results

### Baseline Characteristics of Patients

A total of 102 patients from the safety-net cohort and 145 from the private cohort met the criteria of medically refractory epilepsy and underwent surgical treatment. Patients in the safety-net cohort were more likely to have earlier age of seizure onset, onset outside of the US, longer duration of epilepsy and self-describe as African-American or Hispanic and less likely to self-describe as white. There were no differences in the groups with respect to age at surgery, gender, seizure burden, or risk factors. Complete baseline characteristics of patients is provided in **Table 1**.

**Table 1.**
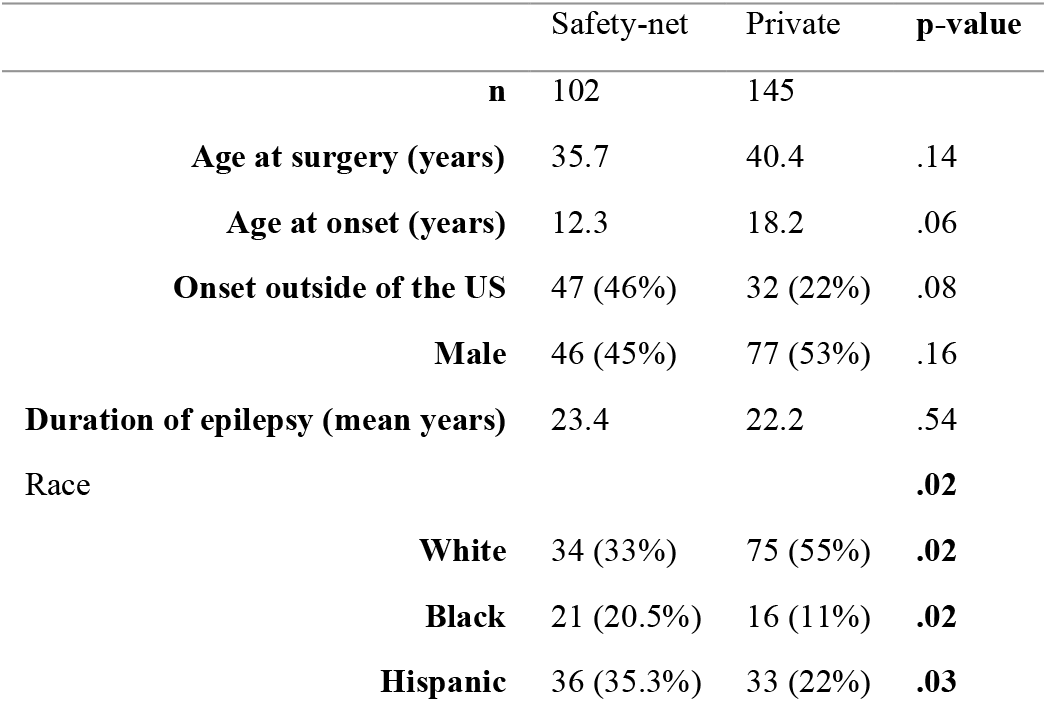
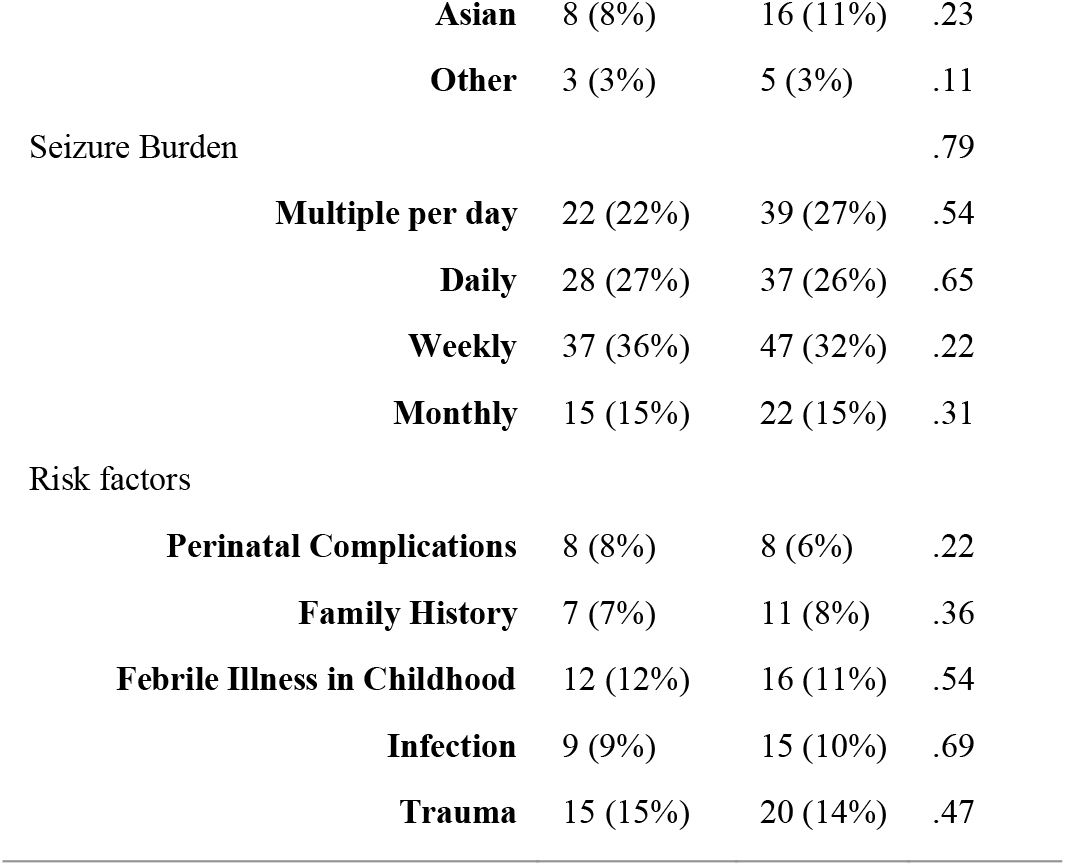
Characteristics of patients receiving surgical epilepsy care at safety-net and private healthcare systems. There was a significant difference in racial groups between safety-net and private healthcare systems (p=.02) but not differences in epilepsy history, seizure burden or risk factors (p>.05)

|  | Safety-net | Private | p-value |
| --- | --- | --- | --- |
| **n** | 102 | 145 |  |
| **Age at surgery (years)** | 35.7 | 40.4 | .14 |
| **Age at onset (years)** | 12.3 | 18.2 | .06 |
| **Onset outside of the US** | 47 (46%) | 32 (22%) | .08 |
| **Male** | 46 (45%) | 77 (53%) | .16 |
| **Duration of epilepsy (mean years)** | 23.4 | 22.2 | .54 |
| Race |  |  | **.02** |
| **White** | 34 (33%) | 75 (55%) | **.02** |
| **Black** | 21 (20.5%) | 16 (11%) | **.02** |
| **Hispanic** | 36 (35.3%) | 33 (22%) | **.03** |

**Table 1. Characteristics of patients receiving surgical epilepsy care at safety-net and private healthcare systems.**
|  |  |  |  |  |
| --- | --- | --- | --- | --- |
|  | **Asian** | 8 (8%) | 16 (11%) | .23 |
|  | **Other** | 3 (3%) | 5 (3%) | .11 |
| Seizure Burden |  |  |  | .79 |
|  | **Multiple per day** | 22 (22%) | 39 (27%) | .54 |
|  | **Daily** | 28 (27%) | 37 (26%) | .65 |
|  | **Weekly** | 37 (36%) | 47 (32%) | .22 |
|  | **Monthly** | 15 (15%) | 22 (15%) | .31 |
| Risk factors |  |  |  |  |
|  | **Perinatal Complications** | 8 (8%) | 8 (6%) | .22 |
|  | **Family History** | 7 (7%) | 11 (8%) | .36 |
|  | **Febrile Illness in Childhood** | 12 (12%) | 16 (11%) | .54 |
|  | **Infection** | 9 (9%) | 15 (10%) | .69 |
|  | **Trauma** | 15 (15%) | 20 (14%) | .47 |

### Comparative Analysis of Presurgical Evaluation

All patients underwent the same standard epilepsy workup including complete seizure history and investigation of prior failed medical therapies, phase 1 monitoring, neuropsychological evaluation, and an epilepsy protocol MRI of the brain. All patients were discussed in the same formal multidisciplinary epilepsy conference with subspecialists from neurology, neurosurgery, radiology, and neuropsychology. A census was agreed upon for any advanced imaging or evaluation (PET, invasive EEG, SPECT, MEG). There were no differences between the groups with respect to the utilization of advanced imaging modalities or phase 2 invasive EEG monitoring (p>.18). However, private hospital patients were more likely to receive MEG imaging (p=.02) as this modality is not available in the safety-net hospital system. A complete list of baseline characteristics of patients is provided in **Table 2**.

**Table 2.**
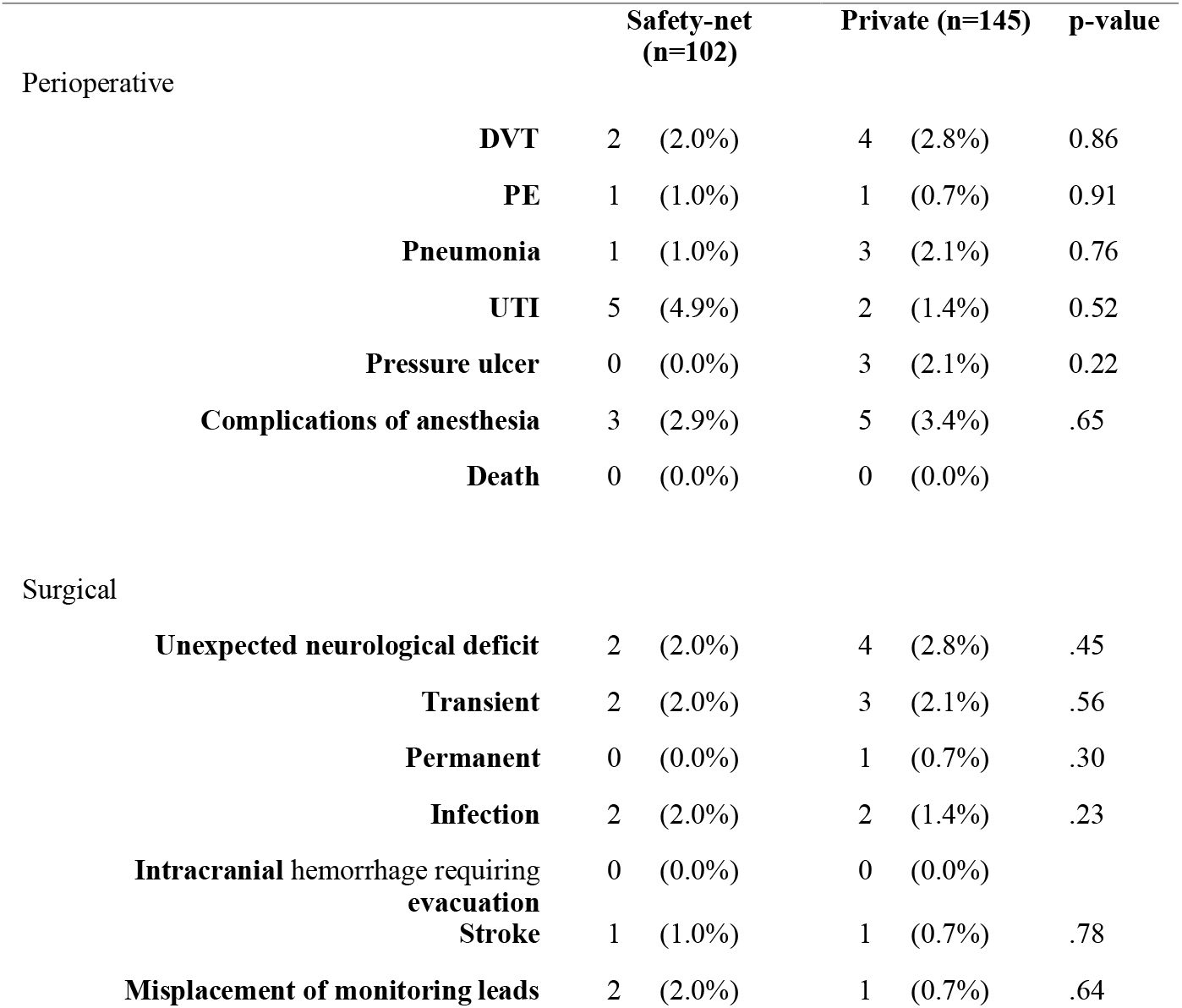
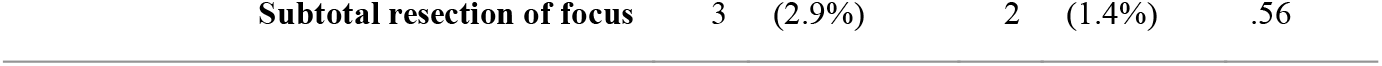
Perioperative and surgical complications. There were no differences in complications between safety-net and private healthcare systems (p>.05)

### Perioperative and Operative Complications

The overall number of perioperative complications in both groups was low. There were no statistically significant differences in the two groups with respective to perioperative complications such as DVT, PE, pneumonia, UTI, pressure ulcer, complications of anesthesia, or death (p>.05). The overall surgical complication in both groups was low. There were no statistically significant differences in the two groups with respect to surgical complications such as neurological deficit, infection, intracranial hemorrhage, stroke, cranial nerve palsy, misplacement of electrodes, or subtotal resection of seizure focus (p>.05). A summary of peri-operative and operative complications and surgical procedures is provided in **Table 3**.

### Surgical outcomes

Engel class 1 outcomes was achieved in 79% and 77% in the safety-net and private hospital groups respectively. Engel class II outcomes was achieved in 16% and 17% in the safety-net and private hospital groups, respectively. Engel class III or IV was observed in a total of 4% in the safety-net group and in approximately 6% of the private group. There was no statistically significant difference in seizure free outcomes between groups (p=.95). However, patients in the safety-net group had a shorter average follow up period as compared to the private group (4.1 years vs. 6.6 years; p=.04). A summary of surgical outcomes is shown in **Figure 1**.

**Figure 1.**
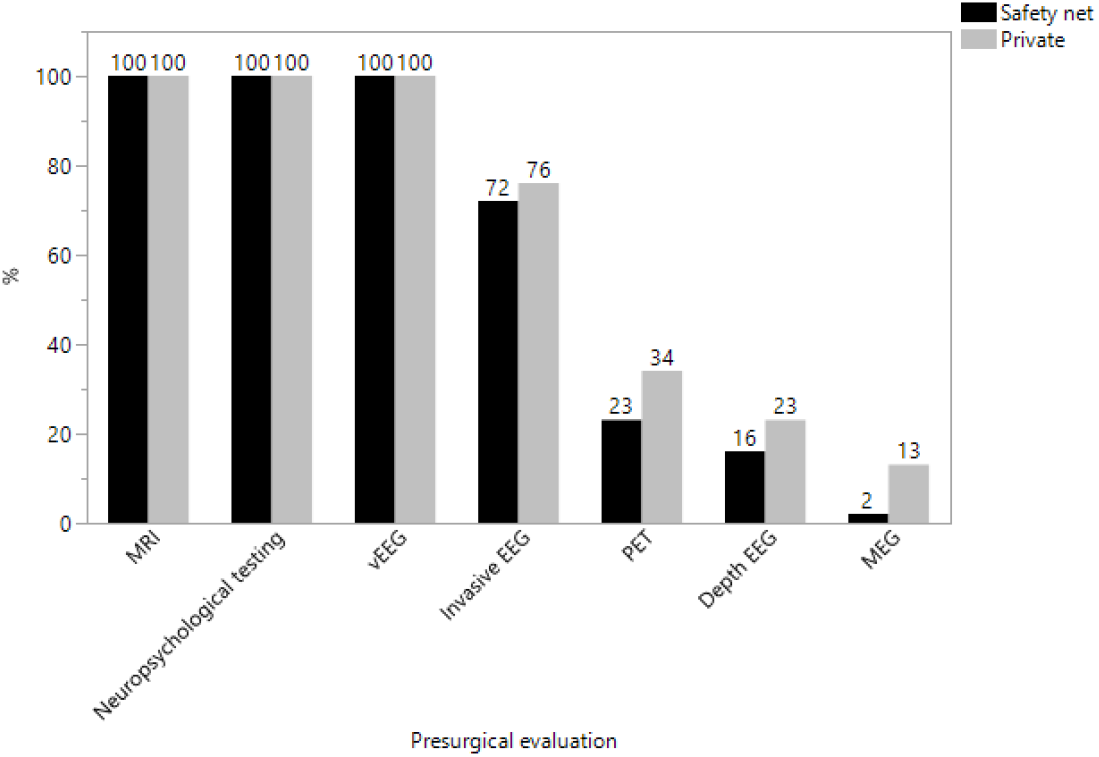
Presurgical evaluation. There were no significant differences in presurgical evaluation between safety-net and private healthcare systems (p>.05) except for MEG (p=.02). Note: the one patient in the safety-net group had previous MEG evaluation outside of safety-net hospital system.

**Figure 2.**
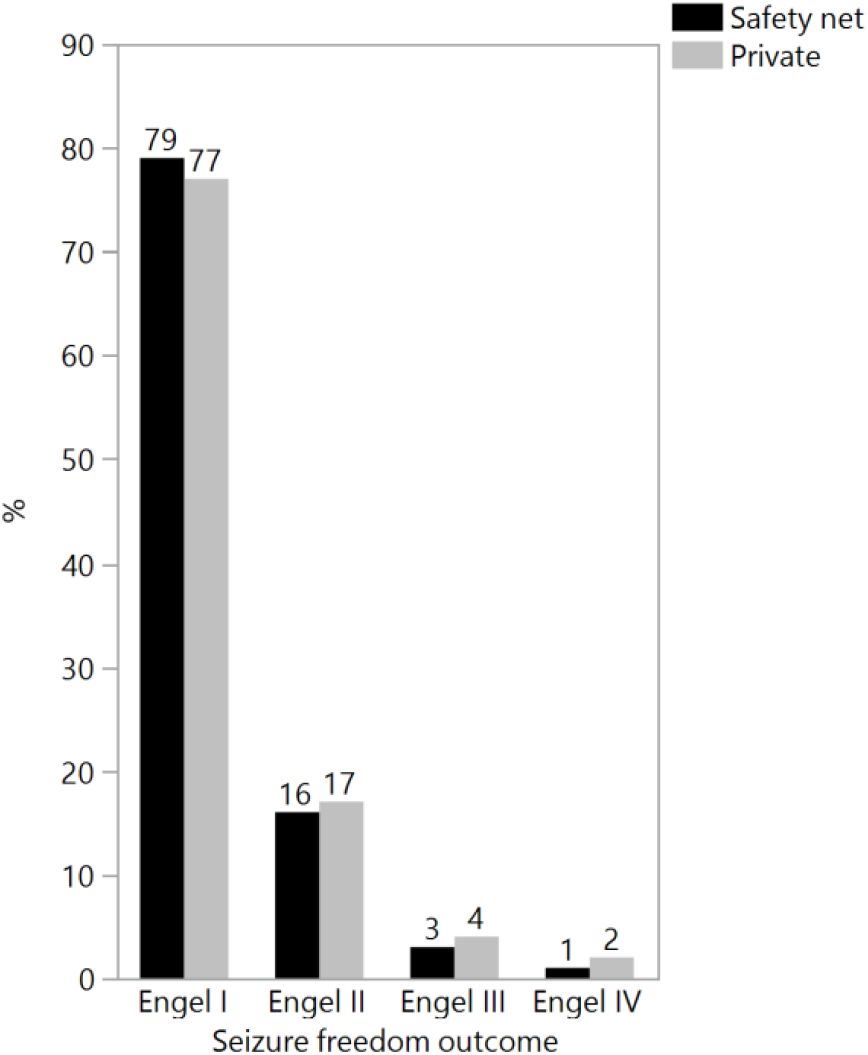
Seizures freedom outcomes. There were no significant differences in seizure freedom outcomes between safety-net and private healthcare systems (p=.95).

## Discussion

To date there have been no comparative analyses of the outcomes of surgical epilepsy care in safety-net versus private hospital systems. Recent reports on the quality of surgical care and costs delivered at safety-net hospital systems raise questions about the delivery of surgical epilepsy care in this setting. Safety-net hospital systems are major providers of health care to patients without insurance and medically and socially vulnerable populations and are often providers of last resort. Patients with medically refractory epilepsy who may benefit the most from surgical care are often those from socioeconomically underserved and racially underrepresented backgrounds who are more likely to utilize safety-net hospital systems for their care.

In an analysis of 247 total patients (102 from safety-net hospital system, 145 from private hospital system) with long term follow up (>4 years), we observed no differences in the postsurgical seizure freedom or control outcomes between the two groups. The overall complication rate was low in both groups with respect to perioperative and operative complications. Patients in both groups received nearly equivalent presurgical evaluation with the exception of higher MEG use in the private hospital system. The safety-net group was represented by a majority of African-Americans and Hispanics compared to a majority of white patients in the private system and on average had a longer duration of epilepsy before surgical intervention.

A number of reports have documented the impact of socioeconomic, racial, and others factors on the delivery and outcomes of surgical epilepsy care. Studies have shown that African-Americans are more likely to have seizure recurrence following epilepsy surgery as compared to other racial and ethnic groups (12). Both African-Americans and Asian or Pacific Islanders with associated limited English proficiency have a significantly longer time to epilepsy surgery after pre-surgical evaluation compared to other racial and ethnic groups (13). African-Americans and the elderly are overall less likely to receive epilepsy surgery if they don’t have private insurance (14). While many of these trends can be attributed to access and health insurance coverage status, it is clear that universal health care is not a comprehensive solution. In a study of a universal health care system with total access and high-quality surgical epilepsy care, less than 2% of eligible patients received epilepsy surgery and 10% of eligible patients who did not died within 2 years (15). However, reports from resource limited environments are encouraging and have highlighted the ability to achieve high rates of seizure freedom with epilepsy surgery and numerous lessons can be learned from their implementation and outcomes. In reports from India and Chile, authors have described the resource limited environments in which surgical epilepsy care has been provided with rates of seizure freedom that parallel what has been reported in the United States at a fraction of the price (16-17). Indeed, there is tremendous opportunity to provide surgical epilepsy care in the underserved.

Safety-net hospitals are likely to play a significant and increasing role despite the reduction in the number of uninsured individuals. In Massachusetts, the passage of health care reform was associated with a significant reduction in the number of uninsured patients but also an increased utilization of safety-net hospitals (18). From this data, we anticipate that the millions of newly insured patients will prefer to stay within safety-net hospital systems across the entire spectrum of care.

Comprehensive epilepsy care is complex, requiring the resources of a multidisciplinary team with significant experience. The two hospitals in our study were part of the USC Epilepsy Care Consortium that spans six independent institutions, both public and private. Management of surgical epilepsy patients, from prediagnostic workup to surgical plan, are discussed in a weekly multidisciplinary multi-institutional meeting. This enables pooling of expertise and standardization of management across different healthcare settings. On the other hand, the partnership of the safety-net hospital analyzed in this study with a private hospital system limits the generalizability of this data to other safety-net hospitals without such a partnership. We interpret this to mean that with proper partnership and support, quality surgical epilepsy care can be delivered in a safety-net hospital system. Intrinsic limitations or qualities of safety-net hospital systems, however, can be overcome in the right circumstances.

Although data was collected in a prospective fashion, there was no true treatment control and therefore, the analysis is observational in nature. In addition, the analysis is limited to patients who underwent resective surgery for epilepsy and does not include patients who are primarily medically managed or who undergo alternatives to resection such as neuromodulation with responsive neurostimulation, vagal nerve stimulation, deep brain stimulation, or laser ablation. However, as the complexity of those procedures is not significantly different from the treatment studied in this group, we would not expect the results to be dissimilar between the safety-net and private hospital systems.

In conclusion, we have shown that safety-net hospitals can provide epilepsy care with equivalent rates of seizure freedom and comparably low morbidity and mortality as private hospital systems. This occurs in the context of treating an population that has been shown to be historically predisposed to poorer quality of care. Designing and implementing quality surgical epilepsy care programs in this population has its own challenges and rewards and is paramount for modern and equitable epilepsy care in the United States.

## Data Availability

Data produced in the present study area are not shared.

## References

1. Wiebe S., Blume W., Girvin J., Eliasziw M., A Randomized, Controlled Trial of Surgery for Temporal-Lobe Epilepsy. N Engl J Med 2001; 345:311–318 August 2, 2001.

2. Langfitt J. Cost-effectiveness of anterotemporal lobectomy in medically intractable complex partial epilepsy. Epilepsia 1997;38:154–163.

3. Mikati MA, Comair YG, Rahi A. Normalization of quality of life three years after temporal lobectomy: a controlled study. Epilepsia 2006;47:928–933.

4. Leestma JE, Annegers JF, Brodie MJ, et al. Sudden unexplained death in epilepsy: observations from a large clinical development program. Epilepsia 1997; 38: 47–55.

5. Hesdorffer DC, Tomson T, Benn E, et al, and the ILAE Commission on Epidemiology; Subcommission on Mortality. Combined analysis of risk factors for SUDEP. Epilepsia 2011; 52: 1150–59.

6. Sperling MR, Barshow S, Nei M, Asadi-Pooya AA. A reappraisal of mortality after epilepsy surgery. Neurology 2016; 86: 1938–44.

7. Sutton JP, Washington RE, Fingar KR, Elixhauser A. Characteristics of Safety-Net Hospitals, 2014: Statistical Brief #213. Healthcare Cost and Utilization Project (HCUP) Statistical Briefs [Internet]. Rockville (MD): Agency for Healthcare Research and Quality (US); 2006-2016 Oct.

8. Hoehn RS, Wima K, Vestal MA, Weilage DJ, Hanseman DJ, Abbott DE, Shah SA. Effect of Hospital Safety-Net Burden on Cost and Outcomes After Surgery. JAMA Surg. 2016 Feb;151(2):120–8. doi: 10.1001/jamasurg.2015.3209.

9. Lewin M, Altman S. America’s Health Care Safety-net: Intact but Endangered. Washington, DC: National Academy Press; 2000.

10. Mouch CA, Regenbogen SE, Revels SL, Wong SL,Lemak CH, Morris AM. The quality of surgical care in safety-net hospitals: a systematic review. Surgery. 2014;155(5):826–838.

11. Engel, Cascino, Ness, Rasmussen, Ojemann. Outcome with respect to epileptic seizures J. Engel (Ed.), Surgical treatment of the epilepsies, Raven Press, NY (1993)

12. Burneo JG, Knowlton RC, Martin R, Faught RE, Kuzniecky RI Race/ethnicity: a predictor of temporal lobe epilepsy surgery outcome? Epilepsy Behav. 2005 Nov;7(3):486–90.

13. Thompson AC, Ivey SL, Lahiff M, Betjemann JP. Epilepsia. Delays in time to surgery for minorities with temporal lobe epilepsy. 2014 Sep;55(9):1339–46. doi: 10.1111/epi.12700. Epub 2014 Jul 9.

14. McClelland S 3rd, Guo H, Okuyemi KS. Racial disparities in the surgical management of intractable temporal lobe epilepsy in the United States: a population-based analysis. Arch Neurol. 2010 May;67(5):577–83. doi: 10.1001/archneurol.2010.86.

15. Burneo JG, Shariff SZ, Liu K, Leonard S, Saposnik G, Garg AX. Disparities in surgery among patients with intractable epilepsy in a universal health system. Neurology. 2016 Jan 5;86(1):72–8. doi: 10.1212/WNL.0000000000002249. Epub 2015 Dec 7.

16. Rao MB, Radhakrishnan K. Is epilepsy surgery possible in countries with limited resources? Epilepsia. 2000;41 Suppl 4:S31–4.

17. Campos MG, Godoy J, Mesa MT, Torrealba G, Gejman R, Huete I. Temporal lobe epilepsy surgery with limited resources: results and economic considerations. Epilepsia. 2000;41 Suppl 4:S18–21. Review.

18. Ku L, Jones E, Shin P, Byer F, Long S. Safety-net providers after health care reform: lessons from Massachusetts. Archives of Internal Medicine. 2011;171(15):1379–84.

